# Acceptability of diagnostic tampon self-sampling for HPV: Mixed methods findings from the STAMP Trial and focus groups

**DOI:** 10.1101/2025.07.12.25331431

**Authors:** Michelle Gomes, Hannah McCulloch, Valentina Milanova, Kalina Mihaylova, Karin Hellner

**Author notes:** Joint first author - These authors contributed equally to this work (Michelle Gomes, Hannah McCulloch).

## Abstract

**Background:** Multiple barriers limit access to traditional in-person clinician sampling for cervical cancer. The STAMP (Screening with Tampons: Evaluating Diagnostic Accuracy for HPV and Assessing Participant Views) study evaluated the diagnostic accuracy and acceptability of tampon-based self-sampling for HPV amongst people assigned female at birth. Encouraging technical performance of the tampon as a sample for microbial detection was observed relative to clinician-taken samples. Pre and post trial questionnaires and a nested qualitative study explored acceptability.

**Methods:** All participants (n=263) completed questionnaires before and after sampling using three methods: tampon-based self-sampling, swab-based self-sampling, and clinician-collected samples. Questions included comfort, trust and ease of use of tampon-sampling. Four focus groups (n=24) were conducted online, and analysed using reflexive thematic analysis.

**Results:** Questionnaire responses suggested tampon-based self-sampling was highly acceptable. Qualitative analysis identified four key themes: advantages of self-sampling over clinician-sampling; unique benefits of tampon-based self-sampling; concerns about accuracy and trust; and confidence in correct use. Participants valued the physical comfort, psychological ease, and practical convenience of self-sampling compared to clinician-sampling. The familiarity of tampons as an everyday product was central to acceptability, normalising screening. While some initially questioned accuracy, most were reassured by evidence of performance. Clear instructions and understanding the evidence base boosted confidence.

**Conclusion:** Findings suggest that tampon-based self-sampling is a highly acceptable alternative sampling approach for cervical screening that may increase screening participation by addressing multiple barriers. Implementation strategies should emphasise the method’s everyday nature, while providing accessible guidance to support correct usage.

*What is already known on this topic:*

- Self-sampling for Human Papillomavirus (HPV) has been recommended by the World Health Organisation as an additional approach to expand cervical cancer screening services, complementing conventional clinician-taken sampling to broaden access and address barriers associated with in-person care.

*What this study adds:*

- Findings from this study suggest that self-sampling has the potential to support screening for cervical cancer as an acceptable, at-home approach addressing physical, psychological and practical barriers associated with traditional clinician collected sampling.
- Participants reported that the familiarity of using a tampon for self-sampling creates a unique advantage over other self-sampling methods through demedicalising and normalising screening, therefore potentially increasing accessibility and participation among underscreened populations who experience most barriers.

*How this study might affect research, practice or policy:*

- Self-sampling including tampon-based self sampling offers a promising, acceptable approach to achieving cancer elimination goals.
- Implementation of tampon-based self-sampling for HPV should be clear, comprehensive and accessible, incorporating visual aids and simple language while emphasising the rationale and scientific evidence behind the collection method

## Introduction

Cervical cancer remains a significant global health concern despite being largely preventable through effective screening and early intervention (1). In the UK, cervical cancer screening coverage is currently 68.8% among eligible women aged 25-64, leaving a substantial proportion vulnerable (2), despite overwhelming evidence that early detection of high-risk human papillomavirus (HPV) prevents cancer development through timely intervention (3).

Cervical screening inequities disproportionately affect specific populations. Women from ethnic minority backgrounds, particularly South Asian and Black communities, show up to 14% lower coverage than White British women (4). Socioeconomic deprivation strongly associates with reduced participation (5), alongside barriers faced by people with physical or learning disabilities, LGBTQ+ individuals with a cervix, and survivors of sexual violence (6,7).

Traditional speculum-based screening presents multiple participation barriers. Physical barriers include clinic access limitations, appointment availability, and speculum-associated discomfort (8,9). Psychological barriers encompass embarrassment, anxiety, trauma from previous experiences, and concerns about bodily exposure (10). These barriers disproportionately affect underserved populations, contributing to screening inequities and higher cervical cancer rates among disadvantaged groups (11).

Self-sampling for HPV testing has emerged as a promising solution. The World Health Organisation (WHO) recommends HPV self-sampling as a key strategy for achieving 70% screening coverage, recognising its potential to reach underscreened populations (12). Multiple countries have successfully implemented self-sampling programs: the Netherlands doubled self-sampling uptake from 8% to 16% (2017-2020), while Sweden’s Stockholm region achieved a 10 percentage point increase in overall coverage from 75% to 85% (13,14,15). In March 2025, the UK National Screening Committee recommended that cervical screening providers should be allowed to offer HPV self-sampling to under-screened individuals (16).

Evidence supporting self-sampling efficacy is substantial. The YouScreen trial demonstrated that self-sampling could reach an additional million people for cervical screening (17). The HPValidate study found self-collected vaginal samples perform comparably to clinician-collected specimens, with 75% of participants reporting an excellent experience (18). Systematic reviews confirm HPV testing from self-collected vaginal specimens shows high relative sensitivity and specificity compared to clinician-collected samples when PCR assays are used (19).

While various self-sampling devices have been evaluated, research on tampons remains limited despite use by approximately 4.5 million women in the UK (30% of the menstruating population) (20). With larger surface area potentially improving sample collection compared to swabs, previous studies demonstrate tampons can effectively collect samples for HPV detection (21).

The STAMP (Screening with Tampons: Evaluating Diagnostic Accuracy for STIs, BV and HPV and Assessing Participant Views) trial evaluated diagnostic accuracy and acceptability of tampon-based self-sampling compared to traditional methods. Results demonstrated that HPV detection on the diagnostic tampon was highly concordant with clinician-collected samples (22). Essential for accessible and inclusive screening approaches, we explore the acceptability of tampon-based self-sampling for HPV.

## Methods

The STAMP trial assessed diagnostic tampon acceptability via pre- and post-sample questionnaires and a nested qualitative study with online focus groups. Ethical approval was granted by London Camberwell St Giles Ethics Committee (ref: 23-LO-0882). The study enrolled sexually active individuals assigned female at birth (AFAB) aged 25–65 years.

Participants were recruited through social media. Full trial methods are reported elsewhere (22).

A total of 263 women and people AFAB completed acceptability questionnaires before and after self-sampling using vaginal self-swabs (Copan FLOQSwabs) and a diagnostic tampon, alongside clinician-collected sampling. Of those interested, 24 participated in four online focus groups (5–7 per group), most having received HPV, chlamydia, and gonorrhea results prior.

Supplementary materials include the questionnaire and focus group agenda.

Focus groups, facilitated by MG, MT, and VM via Google Meet, lasted approximately 60 minutes, were recorded, transcribed, anonymised, and accuracy-checked by MT, MG, and HM. Participants received £25. Facilitators were female staff from the tampon-developing organisation, unknown to participants, with no prior relationship beyond informed consent.

Questionnaire data were analysed descriptively. Qualitative focus group data were analysed using reflexive thematic analysis (23). HM and MG reflected on positionality before independently coding two transcripts using Dedoose software, then collaboratively developed coding frameworks and initial themes. HM re-coded all transcripts using the refined framework. MG triangulated thematic summaries against field notes and facilitation experience. The final coding framework was agreed following review and refinement with all authors (see supplementary material). Our reporting follows the Consolidated criteria for reporting qualitative research (COREQ) framework.

### Public Patient Involvement and Engagement (PPIE)

The STAMP study design and tampon instructions were reviewed by NHS clinicians and user tested with the public. No patients or members of the public were involved in setting the research question or outcome measures, nor were they involved in developing plans for recruitment, design, or implementation of the study.

## Results

Participant characteristics are shown in table 1. Both the full questionnaire sample and the focus group sub-sample represented diverse demographic backgrounds.

**Table 1:** Questionnaire and focus group participant characteristics.

|  | Questionnaire sample,<br>n=263 (n, %) | Focus group sample,<br>n=24 (n, %) |
| --- | --- | --- |
| Age<sup>1</sup> | 31 (28, 36) | 34 (29, 39) |
| Ethnicity |  |  |
| Asian | 16 (6.1) | 2 (8.3) |
| Black, Caribbean or African | 25 (9.5) | 3 (12.5) |
| Other/Mixed ethnicity | 39 (14.8) | 3 (12.5) |
| White | 183 (69.6) | 16 (66.7) |
| Sexual Orientation |  |  |
| Heterosexual | 197 (74.9) | 20 (83.3) |
| Bisexual | 49 (18.6) | 3 (12.5) |
| Prefer not to say | 7 (2.7) | - |
| Lesbian/Gay | 6 (2.3) | 1 (4.2) |
| Other | 4 (1.5) | - |
| Relationship Status |  |  |
| Single | 155 (58.9) | 16 (66.7) |
| Co-habiting | 53 (20.2) | 6 (25.0) |
| Married | 27 (10.3) | 2 (8.3) |
| Civil Partnership | 12 (4.6) | - |
| Prefer not to say | 10 (3.8) | - |
| Divorced | 6 (2.3) | - |

<sup>1</sup>Median (IQR); n (%)

### Questionnaire results

Almost all participants had prior experience of tampons before the trial (98.5%, 259/263), and 70.7% (186/263) of HPV testing (table 2). Comfort levels with tampon use for testing remained high before and after sampling (very comfortable:206/263, 78.3% pre-sampling;192/263, 73.0% post-sampling), and perceived ease of use increased from 63.5% (167/263) to 74.5% (196/263). Moderate or extreme concerns about accuracy slightly increased post-sampling (10.2% [27/263] to 13.6% [36/263], *p* < 0.05). Of all methods used in the trial, tampons were the preferred method (51.0%, 134/263), followed by self-swabs (43.7%, 115/263) and then clinician-taken swabs (5.3%,14/263).

**Table 2:** Questionnaire and focus group participant characteristics.

|  | **Pre-sampling survey,**<br>**n=263 (n, %)** | **Post-sampling survey,**<br>**n=263 (n, %)** |
| --- | --- | --- |
| Tampon use prior to trial | 259 (98.5) |  |
| Previously tested for HPV | 186 (70.7) | - |
| Level of comfort with idea of using a tampon for sample collection |  |  |
| Very comfortable | 206 (78.3) | 192 (73.0) |
| Comfortable | 51 (19.4) | 49 (18.6) |
| Neither comfortable nor uncomfortable | 6 (2.3) | 5 (1.9) |
| Uncomfortable | - | 5 (1.9) |
| Very uncomfortable | - | 12 (4.5) |
| Perceived ease of tampon use for sample collection |  |  |
| Very Easy | 167 (63.5) | 196 (74.5) |
| Easy | 84 (31.9) | 54 (20.5) |
| Neither easy nor difficult | 7 (2.7) | 8 (3.0) |
| Difficult | - | 3 (1.1) |
| Very difficult | 5 (1.9) | 2 (0.7) |
| Concerns about accuracy when using a tampon for sample collection |  |  |
| Not at all concerned | 69 (26.2) | 63 (23.9) |
| Slightly concerned | 98 (37.3) | 115 (43.7) |
| Somewhat concerned | 69 (26.2) | 49 (18.6) |
| Moderately concerned | 19 (7.2) | 25 (9.5) |
| Extremely concerned | 8 (3.0) | 11 (4.1) |
| Trust in tampon results compared to clinician swabs |  |  |
| Equal trust in tampon and clinician swabs | 151 (57.4) | 144 (54.7) |
| Greater trust in clinician swab | 106 (40.3) | 114 (43.3) |
| Greater trust in tampon | 6 (2.3) | 5 (1.9) |
| Of the methods used as part of this trial, which is your preferred method? |  |  |
| Tampon |  | 134 (51.0) |
| Self-swab |  | 115 (43.7) |
| Clinician swab |  | 14 (5.3) |
| If tampon sample collection were proven effective, I would consider using it for regular screening: |  |  |
| Agree | 254 (96.6) | 251 (95.4) |
| Neither agree or disagree | 7 (2.7) | 10 (3.8) |
| Disagree | 2 (0.8) | 2 (0.8) |
| If tampon sample collection were proven effective, I would get screened more regularly: |  |  |
| Agree | 226 (85.9) | 216 (82.3) |
| Neither agree or disagree | 32 (12.2) | 41 (15.6) |
| Disagree | 5 (1.9) | 6 (2.3) |

### Qualitative analysis of focus group

We identified four main themes in our analysis. Throughout these themes, the central organising concept was the familiarity of the tampon, albeit newly applied for HPV self-sampling. Table 3 provides exemplar quotes from the first two themes.

**Table 3.** Themes and quotes.

| Theme | Quotes |
| --- | --- |
| The advantages of self-sampling for HPV vs in-clinic testing | *“It’s such a pain [Going to have a smear test]. I’ve gotta get a bus. I don’t really know where we are. And then it was stupid thing about what should I wear? ...It’s that kind of, like, added level of stress and anxiety and I sat downstairs [in the clinic] going, oh, this is horrible. What if it hurts? What if you know, all these kind of things. And when I did the swab and a tampon at home, I had none of those kind of concerns.”* Focus group B, P5 |
|  | *“As I mentioned, that lady was lovely and put me at ease, but it’s still quite uncomfortable to go and bare all to someone. And it is kinda an invasion of your personal space as well. So doing it at home was so much more comfortable. And just to go back to, like, the accessibility point I made at the beginning, like, you don’t have to take time out of your day to travel. You can do it around. I mean, if you work from home, which I do, you can do it around work. There’s really nothing stopping you from doing it. Like, you don’t have to pay for transport to get anywhere. So Yeah. Just a lot more comfortable, on quite a few levels”.* Focus group B, P1 |
|  | *“I think for me, personally, having to go for, like, smear test, it’s always something that’s really you know, it’s uncomfortable. It’s painful. It’s unpleasant. Like, I do it because it’s important. But it’s never, like, a fun experience. So, you know, if you can kind of do it at home, safely, effectively, and a bit more often. I think that probably a really good thing. It will encourage more people to do it ”* Focus group B, P6 |
| The potential of the diagnostic tampon for increasing options in self-sampling | *“I was just going to say the swab makes it feel like it’s a little bit more of a medical procedure. But everyone should also be normalised to doing that in other ways, what covid tests and everything else so it shouldn’t be too kind of impactful to them in that sense...I think doing the tampon one kind of normalises it a little bit more...because I know we’ve got a few in the group here who don’t use tampons. I tend to use a variety of things ...But I think it makes it [screening] feel a little bit more every day and a little bit less daunting.”* Focus group C, P3 |
|  | *“I’ve used a tampon so many times, I just felt more secure and already know what I’m doing, rather than a self-swab...I think for me the tampon was the easiest method and, like, the most familiar as well.. I found the tampon comfier than the actual, like, speculum. I think it’s just like a familiar kind of, like, feeling to, like, put the tampon up. Versus, you know, you feel quite exposed going into, like, the clinic and but in terms of, like, comfort, ...just once it’s kinda, like, up there, you just forget about it, set a timer. Yeah. Pretty easy. “* Focus group A, P2 |
|  | *“For me, both [the tampon and the self-swab] were easy. It’s also true that I was already used to the swab... I can’t remember, but I think that the swab was hard when I started self-testing with swab....So yeah, the tampon is as easy as the swab for me, but I imagine that for people who has never who have never self-tested, the tampon could be easier. Focus group A, P5* |
|  | *“Facilitator: So for those of you who don’t regularly use tampons, would you say that the fact that it was a tampon as the self-collection method did that give you any kind of anxiety or...apprehension about using the service?*<br>*Participant 5: yeah, it definitely did.... it did make me a bit apprehensive about going through with it. But again, it’s changed my whole perception. So now if I’m in a situation where I’m my friend and I have an accident or I start my period and she says do you want to use this tampon? I definitely take it now, easy. I used to like refuse it and just go run to the shop to get a pad,...but now if I’m that situation, I definitely take a tampon offered”,* Focus group C |

**Table 4.** Barriers and enablers to implementation: Themes, quotes and recommendations.

| Theme | Quotes | Recommendations |
| --- | --- | --- |
| Accuracy and trust in results: reassurance needed for some | "I feel like there's something like being with a medical professional, I feel a bit more at ease, naturally. I feel like I leave it in their hands, like there's less room for errors " Focus group C, P5 | Clear communication of evidence supporting use of tampon-based self-sampling to reassure users of the accuracy of this novel application of a familiar technology |
|  | "Personally, I would have previously trusted the clinicians over my own, but I've had a couple where they've had to repeat the smear cause they didn't get enough cells, and that's a whole faff twice. So after that, I would say my opinion has changed....after those sorts of occurrences I would trust it equally. And I think definitely if you did have data, because I think a lot of people's concerns would be about it comparing, so if you have the data to prove it, then I dont see why not" Focus group D, P3 |  |
|  | "If I could see evidence to show that the correct cells were definitely being examined once it was sent to the lab then it would definitely increase kind of my confidence and I'd be more likely to use it.... it would definitely make a difference but I think without it I would definitely be doubtful" Focus group D, P2 |  |
|  | "I was just thinking that, like, because the tampon thing is new, I wonder if some of us will wonder if it's gonna be as reliable just because it's new. Whereas we all know that, like...self swabbing is newer, but it's something that it sounds like most of us are familiar with. But that, like, for me, if it's in use and the statistics back it up, I would be just as likely to trust it as the old school methods. Because it seems like a tampon has more surface area" Focus Group A, P1 |  |
| Confidence in correct use: guidance not always needed, but welcomed despite familiarity | "I guess it just very much felt like using a normal tampon. So, there's no kind of worry about, like, how far it has to go in or something because it's just something you're used to" Focus group A, P4 | Comprehensive, accessible guidance to support correct use, including visual aids.<br>Procedural guidance should underscore rationale behind insertion procedures and applicator use, ie to ensure correct placement and avoid contamination<br>Provision of spare tampons in screening packs to mitigate against contamination through menstrual spotting |
|  | "I guess it's sort of along the same lines as my doubts on self test [swab] of just like the tampons not going to be as far up inside. Is it collecting cells from the right place? Especially if it's only for 20 minutes...it's kind of only going to get cells from sort of quite near the opening. So yeah again, but that's going back to kind of what I first learned about smear tests many years ago" Focus group C, P2 |  |
|  | "Because this was the first time I'd ever use the tampon I think, it was an easy method to use obviously, we have guidance on tampons that's everywhere anyway,...but I think having the guide to kind of explain...how to use it...yeah, I've felt a bit more sure that I have kind of done it right myself" Focus group C, P4 |  |
|  | "I thought that having an applicator tampon, it probably helped with making sure it went to the right place rather than kind of picking anything up on the way." Focus group C, P2 |  |
|  | "[Correct placement of the tampon near the cervix] probably wasn't a concern of ...didn't really think twice about that because I didn't realise that for HPV, that was kind of, like, really needed. Not something I really thought about. But then also knowing that now if you're using an applicator anyway and that's it's designed sort of, like, get it in the right place, then I think I'll trust that process" Focus group B, P1 |  |
|  | "I think for me the tampon was the easiest method and or, like, the most familiar as well. I really like the fact that they were, like, 2 kind of, like, tampons, as well because I tried the first one, and I noticed that there was, like, a tiny bit of blood. So I was like, okay. I'm gonna wait for a few days" Focus group A, P2 |  |

#### The advantages of self-sampling for HPV vs traditional clinician-collected samples

Participants described a range of physical, psychological, and practical advantages of self-sampling for HPV compared to clinician-collected samples. Many highlighted how self-sampling addresses barriers linked to clinic-based smear tests.

Physical comfort was a key benefit; self-sampling was described as less invasive and painful than smear tests. Where concerns about discomfort were raised, these were often dispelled after use or outweighed by other advantages. Psychological benefits were particularly valued, participants reported feeling less embarrassed, anxious, and vulnerable when testing at home. Several found traditional smear tests emotionally distressing. One participant stated:

> *“I hate the smear… the whole thing makes me feel sick and slightly violated. I’m not sure if it’s like personal anatomy or whatever, but I find it’s really actually like borderline brings me to tears … the nurses are always lovely, don’t get me wrong. It’s never them. It’s just the whole process”* Focus group D, P3

In contrast, the autonomy of self-sampling was highly valued. Participants appreciated having control over the process in their own space and at their own pace. This contrasted sharply with the perceived loss of control in examinations.

Practical benefits, especially the convenience of home testing, were frequently cited. Participants appreciated avoiding appointment scheduling, travel, and time away from work or caregiving. While many acknowledged positive experiences with clinicians, there was a strong preference for self-sampling overall. Participants believed its combined benefits could help increase screening uptake among those deterred by clinic-based barriers.

#### The potential of the diagnostic tampon for increasing options in HPV self-sampling

Using a tampon as a self-sampling device was seen to offer all the physical, psychological, and practical benefits associated with self-sampling more broadly. In addition, almost all participants appreciated how familiar and easy the method felt. This familiarity as an everyday item helped normalise and demedicalise screening, making it feel less clinical, more routine, and even empowering. It reduced psychological barriers, allowing screening to fit more seamlessly into everyday life. Swabs were familiar to some participants, through testing in a sexual health or COVID context, but were not framed or demedicalised as an everyday item.

> *“When it doesn’t feel as kind of serious in that sense,… you’re doing your own medical treatment*… *[so] If it became an everyday item like a tampon, I don’t think anyone would bat an eyelid at it. I think they’d be more inclined to use it and feel far more relaxed about it…*.*”* Focus group C, P4

Participants identified groups they felt could especially benefit from tampon-based self-sampling, such as those late or reluctant to screen, young people, and individuals with a history of sexual assault. One participant shared they had not attended screening in 10 years but would consider this method. Two participants who did not regularly use tampons welcomed it as an option, despite some initial apprehension due to “*the unfamiliarity*”. While both self-sampling methods were positively received, some expressed a preference for self-swabs, whilst others expressed a preference for tampons.

#### Barriers and enablers to use of the diagnostic tampon for screening

The final themes relate to barriers and enablers to implementation of the tampon as a self-sampling option for HPV testing. See table 3 for exemplar quotes and subsequent recommendations for implementation.

#### Accuracy and trust in results: reassurance needed for some

Participants shared preconceptions about the accuracy of the tampon as a novel self-sampling device. Many assumed that clinician-collected samples would be the more accurate sampling approach, based on clinicians’ medical training and experience. Others held contrasting views, informed by past experiences of inconclusive or incorrect results from clinician-led sampling.

When informed that tampon-based self-sampling has high diagnostic accuracy for detecting high-risk HPVs, most participants felt reassured and more confident. This evidence often shifted their perspective, although a small minority still preferred clinician-collected tests.

> “I think generally speaking, I sort of had this idea that the test done by the medical professional would be, like, more accurate. But, you know, to know that the results are just as good, just as valid, as long as you do it properly at home, is definitely reassuring.” *Focus group B, P6*

Participants stressed the need to clearly communicate the evidence supporting the tampon’s accuracy to potential users. They felt that addressing assumptions about clinician superiority would require transparent, accessible information about validation studies. This was seen as critical to building trust in the method, particularly for those skeptical of self-sampling. Clear communication about the evidence behind the tampon’s performance was viewed as a necessary step toward wider acceptance of this familiar but newly applied technology.

#### Confidence in correct use: guidance not always needed, but welcomed despite familiarity

Participants’ confidence in using tampons for self-sampling was often shaped by its familiarity and procedural simplicity. Many described the process as intuitive and easy, even among those who had never used tampons. Some felt confident in their tampon-sampling technique despite limited or no use of instructions.

However, not everyone felt entirely confident with tampon-sampling. Concerns were raised about correct placement (a concern similarly voiced about self-swabs), how long to leave the tampon in, and risk of contamination, especially from menstrual spotting or tampon-handling. These uncertainties sometimes led to reduced trust in accuracy. In one group, a participant who initially felt confident later doubted their technique after hearing others express confusion about whether the tampon needed to reach the cervix. This highlighted a broader lack of understanding around cervical screening processes, whether clinician-led or self-administered.

Participants suggested several ways to build confidence. Clear, concise instructions with visual aids were highly valued, especially when they explained both the process and its rationale.

Information addressing knowledge gaps about HPV sampling was particularly helpful. The spare tampon was also appreciated, offering reassurance that an accurate sample could still be collected if unexpected bleeding occurred.

## Discussion

We found that self-sampling using swabs and tampons represents an acceptable HPV testing approach that addresses multiple barriers to traditional clinician-sampling. In this study, participants valued the physical comfort, psychological ease, and practical convenience of tampons compared to clinician-administered examinations. The familiarity of tampons was a powerful mediator of acceptability, effectively transforming a medical procedure into a routine activity. While some participants initially questioned tampon accuracy, most were reassured by evidence of diagnostic performance.

These findings contribute to growing evidence on self-sampling acceptability (24). While existing literature establishes self-sampling as an acceptable alternative that increases screening participation, few studies have assessed tampons as sampling devices (13, 17, 25, 26). Our participants overwhelmingly preferred self-sampling methods, tampons (51.0%) or swabs (43.7%), over clinician-collected sampling (5.3%). Qualitative findings revealed that tampon familiarity created confidence and competence in self-sampling. Tampon familiarity as an everyday menstrual product demedicalised the sampling process, compared to swabs or brushes, which, for some, even though known through other testing settings, evoked clinical associations and usage uncertainty. This normalisation effect represents a novel, previously unexplored pathway to reducing screening barriers.

The diagnostic tampon appears promising for reaching underscreened populations. Participants who previously avoided or delayed screening due to negative experiences expressed willingness to use tampon-based self-sampling, suggesting potential for addressing screening barriers and inequities among those with trauma histories, anxiety disorders, or cultural barriers to intimate examinations, consistently underscreened groups (4). Australian cervical screening programme insights highlight how innovative self-collection approaches offer more accessible, inclusive, convenient care for underscreened populations while emphasising that one size does not fit all (27).

Several implementation considerations emerged from our analysis. Consistent with recent qualitative self-sampling research, participants expressed concerns about accuracy and self-efficacy (28). Clear communication about self-sampling accuracy appears crucial for building trust. Instructions should balance comprehensive guidance with accessibility, incorporating visual aids and simple language to accommodate diverse literacy levels and learning preferences. Providing spare sampling devices emerged as important for addressing contamination concerns. Research on sexual and reproductive health self-care innovations emphasises the importance of clear communication and wrap-around support when redistributing work from health practitioners to users (29,30).

The study has some limitations. Almost all participants had previously used tampons (98.5%), significantly greater than the estimated proportion of tampon-users in the UK (20). Considering this, and participants’ position as self-selected volunteers, their views may not represent those with strong screening or tampon-use aversions. However, focus groups represented diverse experiences, including participants with screening delays and two non-tampon users. Further qualitative research is needed among specific groups including those with cultural or religious tampon objections, sexual assault survivors, and underserved populations such as ethnic minority communities, transgender individuals with cervixes, and those with physical disabilities. Finally, participants’ research setting experiences with extensive support may not reflect real-world implementation challenges.

## Conclusions

Tampon-based self-sampling represents a promising HPV testing approach that addresses multiple screening barriers through its familiar, non-medicalised nature. As healthcare systems globally implement self-sampling programmes, policymakers should prioritise acceptable innovations that enhance screening experience and population-level uptake. Our findings indicate tampon-based collection could significantly improve accessibility and acceptability among underscreened populations, advancing WHO’s cervical cancer elimination goals.

Expanding self-sampling options enhances individual choice and autonomy, reinforcing that diverse, accessible screening modalities are essential for equitable cervical cancer prevention.

## Acknowledgements

This acceptability study was made possible through the valuable contributions of several key partners. We thank Lindus Health for their excellent organisation of the STAMP clinical trial, which provided the foundation for this nested qualitative study. Special thanks to Mia Taicher for their invaluable support in facilitating the focus group discussions that form the core of this research. Most importantly, we extend our deepest gratitude to the 24 participants who generously shared their time, experiences, and perspectives in the focus group discussions, and to all 263 STAMP trial participants who completed the acceptability questionnaires. Their openness and insights have been instrumental in advancing our understanding of tampon-based self-sampling acceptability and will inform future implementation efforts to improve cervical cancer screening and STI testing accessibility.

## Author contributions

MG made substantial contributions to the conception and design of the work, acquisition and analysis of data through facilitating focus group discussions and managing the STAMP trial implementation; drafted the manuscript; provided final approval of the version to be published; and agrees to be accountable for all aspects of the work.

HMcC made substantial contributions to the analysis and interpretation of data for the work; drafted the manuscript; provided final approval of the version to be published; and agrees to be accountable for all aspects of the work.

VM made substantial contributions to the conception and design of the work and acquisition of data through involvement in study design, implementation, and presence at focus group discussions; critically revised the manuscript for important intellectual content; provided final approval of the version to be published; and agrees to be accountable for all aspects of the work.

KM made substantial contributions to the conception and design of the work and acquisition of data through involvement in study design, implementation, and presence at focus group discussions; critically revised the manuscript for important intellectual content; provided final approval of the version to be published; and agrees to be accountable for all aspects of the work.

KH made substantial contributions to the conception and design of the work through advisory role; critically revised the manuscript for important intellectual content; provided final approval of the version to be published; and agrees to be accountable for all aspects of the work.

All authors meet the BMJ authorship criteria and have approved the final manuscript for submission.

### Acronyms

AFAB: Assigned Female at Birth
BV: Bacterial Vaginosis
HPV: Human Papillomavirus
NHS: National Health Service
STAMP: Screening with Tampons: Evaluating Diagnostic Accuracy for STIs, BV and HPV and Assessing Participant Views
WHO: World Health Organisation

## Funding

This study was supported by funding from the European Innovation Council (EIC) awarded to Anne’s Day Ltd (Daye). The EIC had no role in study design, data collection and analysis, decision to publish, or preparation of the manuscript.

### Patient Consent for Publication

All participants provided written informed consent for participation in the STAMP trial, including consent for questionnaire data and focus group discussions to be used for research purposes.

Participants explicitly consented to the data being analysed, disseminated, and published in academic research outputs. Focus group participants provided additional consent for audio recording and transcription of discussions. All data has been anonymised and no individual participants can be identified from the published material.

### Data availability

Both the full study protocol and the study data are available on reasonable request, with requests made to the corresponding author.

### Ethical Approval

Ethical approval was obtained from London Camberwell St Giles Ethics Committee (reference 23-LO-0882).

